# Effect of combining lower- and higher-value monthly cash transfers with nutrition-sensitive agriculture, male engagement, and psychosocial intervention on maternal depressive symptoms in rural Malawi: a secondary analysis of a cluster-randomised controlled trial

**DOI:** 10.1101/2024.12.31.24319814

**Authors:** Lilia Bliznashka, Odiche Nwabuikwu, Marilyn Ahun, Natalie Roschnik, Brenda Phiri, Esnatt Gondwe-Matekesa, Monice Kachinjika, Peter Mvula, Alister Munthali, Daniel Maggio, Mangani Katundu, Kenneth Maleta, Melissa Gladstone, Aulo Gelli, the MAZIKO trial team

**Affiliations:** International Food Policy Research Institute, Washington, DC, USA; Global Academy of Agriculture and Food Systems, University of Edinburgh; Department of Medicine, McGill University, Montréal, Canada; Save the Children UK, London, UK; Save the Children Malawi, Lilongwe, Malawi; Give Directly Malawi; Palm Consulting Ltd, Malawi; Rutgers University; University of Malawi, Zomba, Malawi; Kamuzu University of Health Sciences, Blantyre, Malawi; University of Liverpool, Liverpool, UK

## Abstract

Maternal depression affects one in five women in Malawi. Integrated interventions simultaneously addressing multiple risks are a promising strategy to improve mental health. This study evaluated the impact of a nutrition-sensitive social behaviour change (SBC) interventions (agriculture and livelihoods, male engagement, and Caring for the Caregiver) with or without cash transfers on maternal perinatal depression during the lean season in rural Malawi. A midline survey for a cluster-randomised controlled trial was conducted, where 156 clusters were randomly assigned to four arms (39 clusters/arm): (1) Standard of care (SoC), (2) SBC, (3) SBC+low cash (USD17 per month), and (4) SBC+high cash (USD43 per month). Pregnant women and mothers of children <2 years of age (n=2,682) were enrolled at baseline (May-June 2022). A subsample of 1,303 women were followed-up at midline (November-December 2023). Maternal perinatal depression was assessed using the Self-reporting Questionnaire (SRQ-20) with a score of ≥8 indicating symptoms consistent with depression. Intervention effects were estimated using linear mixed effects models. At midline, SBC+high cash reduced depression scores relative to SoC (mean difference (MD) -1.13 (95% CI -1.96, -0.31)) but had no impact on the proportion of women with depressive symptoms. SBC+low cash and SBC alone had no impact on depression scores or the proportion of women with depressive symptoms. Relative to SBC alone, adding cash to SBC reduced depressions scores and the proportion of women with depressive symptoms regardless of the size of the cash transfer. Cash transfers integrated with SBC can benefit maternal perinatal depression health in rural Malawi during the lean season.

**Key messages:** *What is already known on this topic?:*

- Cash transfers can improve mental health by addressing financial and economic risks, but evidence on perinatal mental health is limited, particularly during periods of peak food insecurity.
- Parenting interventions have no overall impact on maternal mental health, but individual interventions and interventions engaging male caregivers have shown positive effects on perinatal mental health.
- Nutrition-sensitive agricultural interventions can reduce depressive symptoms by improving food security, food access, and household income; however, evidence is scarce.
- Evidence on the additive or synergistic effects of cash transfers, nutrition-sensitive agriculture, and parenting interventions on perinatal mental health is lacking.

*What this study adds?:*

- Larger cash transfers with nutrition-sensitive social behaviour change (SBC) interventions comprising nutrition-sensitive agriculture, male engagement, and psychosocial components reduced maternal depressive symptoms during the lean season. Smaller cash transfers with SBC or SBC alone had no impact on perinatal mental health.
- Cash transfers had an added benefit for maternal depressive symptoms on top of SBC regardless of the size of the transfer.

*How this study might affect research, practice or policy?:*

- Comprehensive intervention packages addressing multiple vulnerabilities and risks for mental health and providing a variety of coping and thriving strategies can help improve perinatal mental health, particularly during peak periods of food insecurity.
- Additional research is needed to understand the precise pathways through which integrated intervention packages work to improve perinatal mental health.

## Introduction

Mental disorders are a major public health concern globally and a leading cause of years lived with disability. Depression and anxiety are the two most common mental disorders globally.^1^ Maternal mental disorders, particular perinatal depression which occurs during pregnancy and the first year post-partum, can lead to pregnancy-related morbidity, mortality, adverse birth outcomes, and poor child health, growth, and development.^2–8^ Maternal depression can also adversely affect parenting and caregiving practices throughout infancy and childhood.^8,9^

In Malawi, depression is the second most prevalent mental disorder with the prevalence increasing by 20% over the past decade.^10^ Perinatal depression affects one in five women, with prevalence ranging from 10% to 33%.^11^ Among the primary risk factors for perinatal depression empirically demonstrated in Malawi are low socio-economic status, lack of social support, experience of intimate partner violence, and HIV infection.^12–16^ There is no routine screening for perinatal depression in primary care settings in Malawi.^11^ There is also substantial treatment gap defined by a lack of mental health professionals.^17^ Therefore, improving prevention by addressing key risk factors can be a viable strategy to reduce the burden of perinatal depression in the country.

Given that poverty is a major risk factor for depression,^18^ and that depression and poverty are mutually reinforcing,^1,19^ social protection interventions (i.e., interventions aiming to reduce poverty and increase household resilience to shocks) are a promising strategy to improve mental health. Consistent evidence shows that cash transfers improve mental health with larger effects for larger or unconditional cash transfers.^20–23^ However, much of this evidence is “very low quality”, arising from studies with serious risk of bias.^22^ In Malawi, studies show that the national Social Cash Transfer programme, providing unconditional cash transfers of USD3-7 per month (MWK2600-5200) depending on household size, improved perceived quality of life and subjective well-being.^24–26^ Further, the Malawi Incentive Programme (cash transfers of MWK500-1000 or MWK2000-4000 (USD 0.30-0.6 or USD 1.2-2.3, respectively) provided to individuals/couples conditional on maintaining their HIV status for a year) improved mental health, with larger effects among those with worse baseline mental health.^27^

However, much of the evidence on cash transfers arises from large social protection programmes targeting ultra-poor and labour-constrained households rather than women during the crucial perinatal period. Presently, there is limited evidence on whether cash transfers work to improve perinatal depression. The Janani Suraksha Yojana programme in India reduced depressive symptoms in pregnant women who received cash transfers of INR1000-1400 (USD12-16) conditional on them delivering in a government health facility.^28^ In contrast, in Tanzania, conditional cash transfers for antenatal care and child growth monitoring (TZS10,000 (USD4.30) per antenatal visit and TZS5,000 (USD2.20) per growth monitoring visit) delivered through a responsive stimulation, health and nutrition intervention had no impact on perinatal depressive symptoms.^29^ Further research is needed to better understand the impact of cash transfers on perinatal depression, as well as the effects of combining cash transfers with interventions targeting the non-economic determinants of depression.^22^

In addition to poverty, food security (i.e., adequate access to “sufficient, safe and nutritious food that meets dietary needs and food preferences for an active and healthy life”^30^) is another main determinant of depression.^18^ The relationship between depression and food security is bidirectional, operating through multiple biological and psychosocial pathways.^31–33^ Therefore, interventions effective in improving food security such as nutrition-sensitive agricultural interventions^34–36^ may be a promising strategy to reduce depressive symptoms. Nutrition-sensitive agricultural interventions can also promote mental health by alleviating poverty through increased food access and household income.^34^ Nevertheless, evidence to date is limited to a single study from Tanzania which demonstrated that a nutrition-sensitive agricultural intervention reduced the odds of probable depression in women with young children by 43%, with improvements in food security explaining approximately one-quarter of the effect.^37,38^ Given the multiple social and cultural risk factors for poor mental health,^1,18^ addressing food insecurity alone is likely insufficient to promote mental health. More research is needed on combining interventions addressing food security with interventions targeting other economic and non-economic determinants of depression.

Among interventions targeting non-economic determinants of depression, parenting interventions (i.e., interventions designed to improve parents’ caregiving knowledge, skills, and practices) can help promote and protect mental health by improving parenting and caregiving practices, mother-child interactions, maternal mood, and psychological wellbeing.^1,17,39^ Although theoretically parenting interventions can create a stable environment that supports mental health, consistent empirical evidence shows that alone they are insufficient to improve perinatal depressive symptoms,^40,41^ even when explicitly designed to do so.^42^ Some individual interventions show promise for improving perinatal depression, including those in Tanzania,^29^ Uganda,^43,44^ and Zambia,^45^ as do interventions involving male caregivers.^46^ To date, no studies have examined the impact of parenting interventions on perinatal depressive symptoms in Malawi.^40^

Social protection, parenting, and nutrition-sensitive agricultural interventions can work in additive or synergistic ways to improve perinatal depression by simultaneously addressing multiple risk factors and providing a variety of coping strategies. Nevertheless, evidence remains limited to the study in Tanzania mentioned above which combined cash transfers with a parenting intervention and found no effect on perinatal depressive symptoms. However, the cash transfers provided in this study were relatively small.^29^ Larger cash transfers are likely necessary in resource poor settings to adequately reduce poverty and address economic risk factors. Integrating psychosocial and psychological interventions, which are among the most effective mental health interventions,^47,48^ can help generate further synergies and enhance the effects on mental health of combining social protection, parenting, and nutrition-sensitive interventions.

In this paper, we examined the impact of MAZIKO – a programme combining maternal and child cash transfers with nutrition-sensitive social behaviour change (SBC) interventions, comprising agriculture and livelihoods, male engagement, and psychosocial components – on perinatal depressive symptoms in rural Malawi during the lean season. The lean season, which typically spans from November until the maize harvest begins in March, is a period of peak food insecurity with recurrent acute food insecurity shocks and large declines in food consumption and dietary diversity and quality.^49,50^

## Methods

### Study design

The MAZIKO trial is described in detail elsewhere.^51^ As of the writing of this paper, the trial is ongoing and will be completed in June 2025. Briefly, the MAZIKO trial is a 3-year cluster-randomised controlled trial implemented in 156 clusters in Balaka and Ntcheu districts in the Southern and Central regions of Malawi. Clusters were defined as village groupings or communities in the catchment areas of community-based childcare centres (CBCCs) targeted by the interventions. Clusters were randomly assigned to one of four intervention arms (39 clusters per arm): (1) standard of care (SoC), (2) SBC, which received agriculture and livelihoods, male champions, and Caring for the Caregiver interventions, (3) SBC+low cash, which received USD17 per month in addition to the SBC, and (4) SBC+high cash, which received USD43 per month in addition to the SBC. The trial aims to evaluate the effectiveness and cost-effectiveness of the SBC and cash transfer interventions on health and nutrition outcomes in women and young children. Maternal depressive symptoms are a pre-specified secondary outcome.^51^ The trial involves outcome measurements at baseline, midline (1.5 years after baseline), and endline (3 years after baseline). The midline survey was designed to understand the fidelity, uptake, and quality of intervention implementation, to measure impacts on selected outcomes likely to be affected by large seasonal changes (e.g., diet and expenditures), and to assess short-term impacts. The endline survey will be designed to assess the impacts on all pre-specified outcomes at the end of the 3-year programme.^51^ This paper used data from the baseline and midline surveys to estimate intervention effects during the lean season, approximately 18 months post-baseline. All analyses were pre-specified.^51^ The trial was prospectively registered with ISRCTN, number ISRCTN53055824.

### Study population

The trial included pregnant women aged 15-49 years and mothers/caregivers (aged 15-49 years of age) of children <2 years of age at baseline living in the study clusters. At the cluster level, all women who met these inclusion criteria were eligible to participate in the MAZIKO interventions. Women were randomly selected for enrolment into the trial. First, a household census was conducted in the CBCC catchment areas prior to the baseline. Using data from the census, we constructed lists of eligible households, i.e., those with women aged 15–49 years who self-reported as pregnant and/or were mothers/caregivers of children <2 years of age. Index mother-child dyads were randomly selected for inclusion in the trial. Randomisation was stratified by pregnancy status. A total of 2,682 mother-child dyads were enrolled at baseline. At midline, we prioritised surveying women pregnant at baseline. Women with children <2 years of age at baseline served as replacements for the fixed budget survey.

Trained enumerators provided information about the study. Written informed consent was obtained from adult women or parents/guardians of women <18 years of age. The trial was approved by the institutional review boards of the University of Malawi (protocol number: 02/22/128) and the International Food Policy Research Institute (protocol number: PHND-22-0214).

### Randomisation and masking

Clusters were randomly assigned to one of four intervention arms using a restricted randomisation procedure, described in detail in the study protocol.^51^ Randomisation was stratified by district to allow for the same approximate number of clusters within each intervention arm in each district. Treatment allocation was modelled using village-level variables to maximise balance across the intervention arms. The algorithm tested 5,000 random allocations. The permutation that minimised the R2 was selected.

Given the nature of the interventions, participants and implementers could not be blinded to intervention assignment. The data collection and analysis teams were blinded to intervention allocation. After being finalised, results were unblinded by a non-study researcher.

### Interventions description

The SoC intervention comprised Care Groups which delivered the government-approved curriculum to improve maternal, infant, and child nutrition and early childhood development through home visits, group sessions, and cooking demonstrations. This component was the same across all four arms.

The nutrition-sensitive SBC comprised agriculture and livelihoods, Male Champions, and Caring for the Caregiver interventions. The agriculture and livelihoods intervention provided agriculture inputs and training using CBCCs and community gardens and increased access to Village Savings and Loans Associations (VSLAs). The male champions approach engaged fathers and husbands through group and couple sessions to improve relationships, to improve sharing of decision-making, resources, and chores, and to reduce gender-based violence. The Caring for the Caregiver package, originally developed by UNICEF and adapted for Malawi by the Department of Nutrition and HIV/AIDS and UNICEF, aimed to improve the capacity of Care Group promoters/cluster leaders and male champions to support and promote women’s mental health and emotional wellbeing.

The cash transfer intervention comprised monthly cash transfers to pregnant women and mothers of young children provided through mobile phones linked to bank accounts registered with participants. Cash transfer values were set at the Malawian Kwacha equivalent of USD17 or USD43 per woman per month, at exchange rates set at baseline in June 2022. These values were determined through an analysis of household consumption and cost of diet.^51^ Further details on the MAZIKO interventions are available elsewhere.^51,52^

### Pathways of impact

The MAZIKO interventions addressed multiple social and cultural distal and proximal risk factors for poor mental health.^1,18^ The SoC Care Groups were expected to improve mental health by supporting maternal and child health and nutrition, improving parenting competencies, and providing social support. The Caring for the Caregiver package was expected to support mental health both directly through activities specifically designed for this purpose and indirectly by promoting social networks and reinforcing support structures through Care Groups. The agriculture and livelihood interventions were expected to improve food security and access to nutritious food through provision of agricultural inputs and training, and to reduce financial constraints through improved access to VSLAs. The Male Champions intervention was expected to improve partner support, fathers’ parenting, and sharing of chores and caregiving responsibilities. In addition, the cash transfers can improve mental health by addressing economic and poverty-related risk factors, reducing financial and liquidity constraints, increasing access to healthcare for the mother and child, and increasing resources and resilience to negative shocks. Lastly, all intervention components were designed to increase women’s empowerment and gender equity, which are associated with improved mental health.^1,53–55^

### Study timeline

The baseline survey was conducted in May-June 2022. Cluster randomisation was completed in August 2022. The Care Group and VSLA interventions started in October 2022, and the cash transfers in November 2022. Distribution of agricultural inputs and agricultural training were completed in March 2023 (winter seeds) and November 2023 (rainfed seeds). The Caring for the Caregiver and Male Champions interventions started in September 2023. The midline survey was conducted in November-December 2023.

### Sample size

The primary outcomes of the MAZIKO trial are women’s mean probability of adequacy of micronutrient intake and child development. Cluster and resource availability suggested ∼40 clusters per intervention arm and 20 mother-child dyads per cluster. Power calculations were conducted only for the primary outcomes and suggested a minimum detectable effect size of 0.31 standard deviations (SDs) for mean probability of adequacy and 0.26 SD for child development.^51^

### Data collection

Structured questionnaires were used at baseline and midline to collect data on socio-economic and demographic characteristics and mental health. At midline, information on self-reported participation in the MAZIKO interventions (care groups, agricultural inputs, VSLAs, and cash transfers) since the start of the programme was also collected. Spouses were not interviewed on participation in the Male Champions component. Enumerators were trained over a three-week period prior to the baseline survey. Topics included research ethics, good clinical practice, questionnaire comprehension, interviewing techniques, CAPI software, and standard operating procedures for the study. A two-week refresher training for the same data collection team was conducted prior to the midline survey.

### Outcomes

Mental health was assessed using the Self-Reporting Questionnaire (SRQ-20), which consists of 20 yes/no questions on depression, anxiety, and stress symptoms.^56^ Questions were asked in private, out of sight/earshot from others. The SRQ-20 has been validated in Malawi against clinical diagnosis with a score ≥8 indicating symptoms consistent with major/minor depression.^57^ Internal consistency was high in our sample at baseline (α=0.84) and midline (α=0.87).

### Statistical analysis

We used linear mixed effects models to estimate intent-to-treat intervention effects of the SBC, SBC+low cash, and SBC+high cash intervention arms compared to SoC. Clusters (the unit of randomisation) were included as a random effect. Per protocol, primary analyses were unadjusted.^51^ We also estimated adjusted estimates controlling for district and *a priori* selected baseline covariates, namely woman’s age (<18 vs ≥18 y), education (primary/lower vs secondary/higher), marital status (married vs unmarried), household head status (self vs other), household head gender (female vs male), and household wealth (lowest vs higher quintiles of total household expenditures). All estimates were mean differences (MD) and 95% confidence intervals (CI). Missing data on SRQ-20, the outcome variable, were not imputed. Missing data on covariates were imputed using mean cluster imputation.

Monitoring data revealed that 50 women assigned to cash transfer arms were not receiving cash transfers. As a sensitivity analysis, we re-estimated the primary analyses excluding these women. As a second sensitivity analysis, we estimated the added benefit of the low cash and high cash intervention arms in addition to SBC by comparing the SBC+low cash and SBC+high cash arms to the SBC arm, as well as the added benefit of extra cash by comparing the SBC+high cash to the SBC+low cash intervention arms. Both unadjusted and adjusted models were estimated. We calculated standardised mean difference (SMD) as a measure of effect size by dividing the unadjusted MD by the pooled standard deviation at baseline.

In addition, we examined effect modification by pre-specified baseline factors: child sex (male vs female), pregnancy status (pregnant vs child <2 years of age), and household wealth (lowest one vs highest four quintiles of household expenditures).^51^ Missing data on effect modifiers were imputed using mean cluster imputation. Interactions were considered statistically significant at *p*<0.10. Post-hoc contrast estimation was used to generate effect estimates for each subgroup of the effect modifiers. All analyses were conducted in Stata Version 18.

## Results

### Sample characteristics

The trial enrolled 2,682 mother-child dyads (**Figure 1**). At midline, 1,303 women had mental health data at both baseline and midline. At baseline, women in the analytic sample were on average 25 years old, 78% were married, 22% had some secondary or higher education, and 89% were pregnant (**Table 1**). Their households had four members, on average, and 32% were headed by women. Mental health was poor with 30% of women experiencing symptoms consistent with depression. **Supplemental Table 1** shows the proportion of women experiencing each of the SRQ-20 symptoms by intervention arm and timepoint. At baseline, characteristics were similar across intervention arms with few exceptions. A higher proportion of households in the SoC arm were in the lowest wealth quintile relative to other intervention arms. In addition, a higher proportion of household heads were female in the SBC arm compared to other intervention arms. Accordingly, a lower proportion of women were married in the SBC arm compared to other intervention arms. *Programme participation*

**Figure 1.**
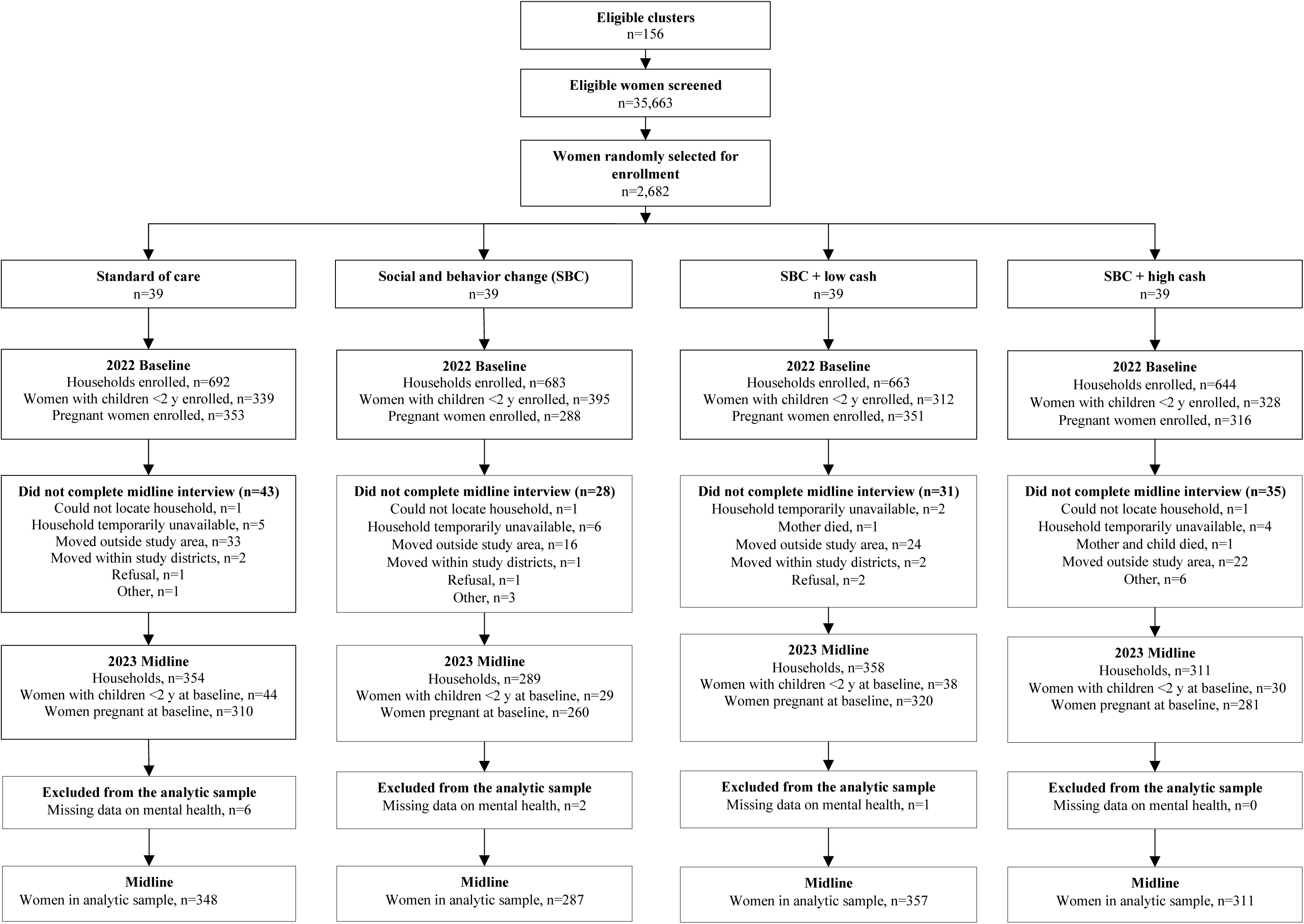
Study flow diagram. Abbreviations used: SBC, social-behavioural change

**Table 1.** Baseline characteristics of the women included in the analytic sample.

|  | **Standard of care** | **Social behaviour**<br>**change (SBC)** | **SBC + Low cash** | **SBC + High cash** |
| --- | --- | --- | --- | --- |
|  | **Mean ± SD or %** | **Mean ± SD or %** | **Mean ± SD or %** | **Mean ± SD or %** |
| N | 348 | 287 | 357 | 311 |
| *Household characteristics* |  |  |  |  |
| Household size | 4.2 ± 1.9 | 4.1 ± 1.8 | 4.1 ± 1.9 | 4.0 ± 1.7 |
| Total household expenditures (MWK) | 35,835 ± 29,256 | 37,264 ± 31,406 | 37,052 ± 29,868 | 36,284 ± 29,487 |
| Lowest household expenditure quintile | 23.3 | 17.1 | 18.2 | 20.9 |
| *Household head characteristics* |  |  |  |  |
| Age (in years) | 33.5 ± 11.6 | 32.0 ± 10.7 | 33.3 ± 11.3 | 33.0 ± 10.8 |
| Head is female | 30.2 | 36.6 | 32.2 | 29.9 |
| Secondary or higher education | 28.5 | 25.8 | 29.4 | 26.4 |
| *Woman's characteristics* |  |  |  |  |
| Age (in years) | 25.3 ± 7.0 | 25.3 ± 7.2 | 25.5 ± 7.1 | 24.8 ± 6.6 |
| Woman is <18 years | 11.5 | 11.2 | 8.7 | 10.6 |
| Pregnant at baseline | 87.1 | 90.2 | 89.3 | 90.8 |
| Married or co-habiting | 81.0 | 76.0 | 78.2 | 77.9 |
| Secondary or higher education | 21.6 | 21.3 | 22.7 | 21.2 |
| Woman is head of household | 22.7 | 25.4 | 23.8 | 18.3 |
| Self-reporting Questionnaire (SRQ-20)<br>score | 5.6 ± 4.2 | 5.8 ± 4.2 | 5.5 ± 4.3 | 5.6 ± 4.1 |
| Has symptoms consistent with<br>depression (SRQ-20≥8) | 31.3 | 28.6 | 29.1 | 30.2 |
| *Child characteristics* |  |  |  |  |
| Child is a boy | 50.0 | 51.1 | 49.8 | 44.4 |
| Child age (months) | 15.4 ± 5.7 | 15.5 ± 5.3 | 15.8 ± 5.1 | 15.4 ± 5.1 |

Self-reported programme participation indicated that Care Group membership was low, ranging from 17% in the SoC arm to 45% in the SBC+high cash arm. In terms of the SBC interventions, 64% of women in the SBC and SBC+low cash arms and 74% in the SBC+high cash arm reported receiving agricultural inputs in the past 12 months. VSLAs membership ranged from 36% in the SoC arm to 48% in the SBC+low cash arm. With respect to cash transfers, 58% of women in the SBC+low cash arm and 68% in the SBC+high cash arm reported being registered in a cash transfer program. Notably, monitoring data showed that only 50 women (4% of the analytic sample) were not receiving cash transfers. A handful of women in the SoC arm reported being registered in any cash transfer program (3%) or receiving agricultural inputs (3%). Likewise, 5% of women in the SBC arm reported being registered in any cash transfer program, likely reflecting receipt of the Malawi Social Cash Transfer program.

### Programme impact

The SBC+high cash intervention significantly reduced SRQ-20 scores at midline by MD -1.13 (95% CI -1.96, -0.31) as compared to SoC, but had no impact on the proportion of women experiencing depressive symptoms (**Figure 2A and 2B, Supplemental Table 2**). We observed a shift in the distribution of SRQ-20 scores such that the SBC+high cash intervention benefited all women (**Supplemental Figure 1**). The SBC and SBC+low cash interventions had no effect on SRQ-20 scores or the proportion of women experiencing depressive symptoms relative to SoC. Intervention effects were similar after adjusting for covariates, albeit point estimates were smaller (**Supplemental Tables 3**). Results from the sensitivity analyses excluding the 50 women who were not registered to receive cash transfers were also consistent (**Supplemental Table 4**).

**Figure 2.**
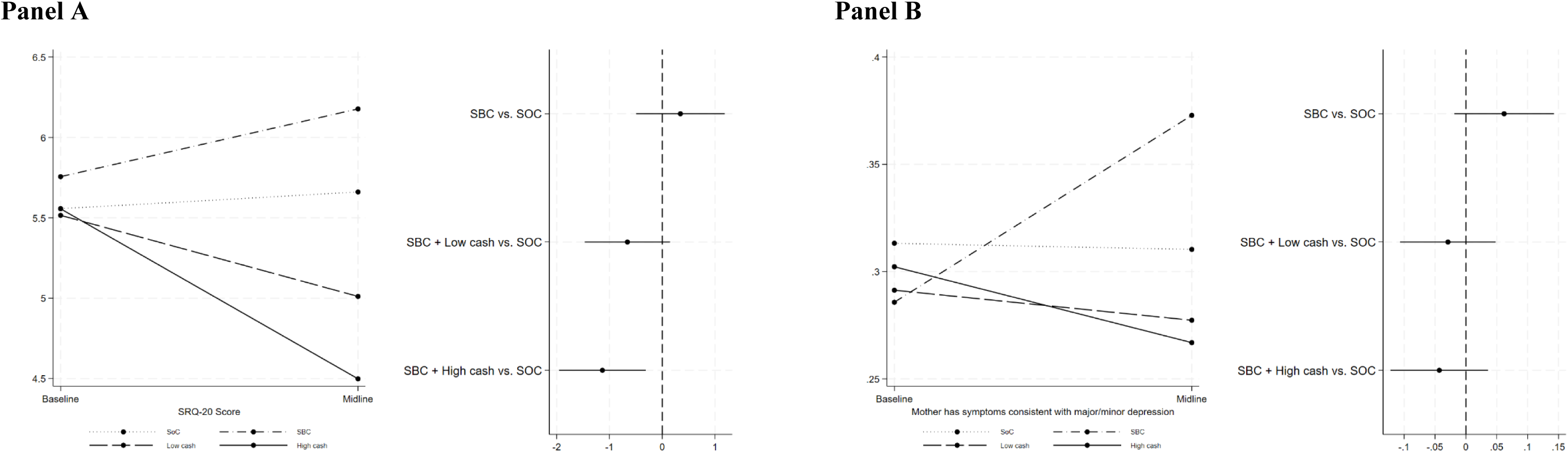
Effect of the MAZIKO interventions relative to the standard of care on depression scores (Panel A) and the proportion of women with depressive symptoms (Panel B). Abbreviations used: SBC, social behaviour change; SOC, standard of care.

The sensitivity analyses examining the added benefit of the cash transfer compared to SBC alone showed that both SBC+low cash and SBC+high cash reduced SRQ-20 scores relative to SBC: MD -1.00 (95% CI -1.84, -0.17) and MD -1.48 (95% CI -2.33, -0.63), respectively (**Figure 3A**, **Supplemental Table 5**). Likewise, SBC+low cash and SBC+high cash reduced the proportion of women with depressive symptoms by 9 and 11 percentage points relative to SBC, respectively (**Figure 3B**, Supplemental Table 5). When comparing SBC+high cash and SBC+low cash, there was no added benefit of the higher cash transfer on SRQ-20 scores or the proportion of women with depressive symptoms (Figure 3, Supplemental Table 5). Results were consistent after adjusting for baseline covariates (**Supplemental Table 6**).

**Figure 3.**
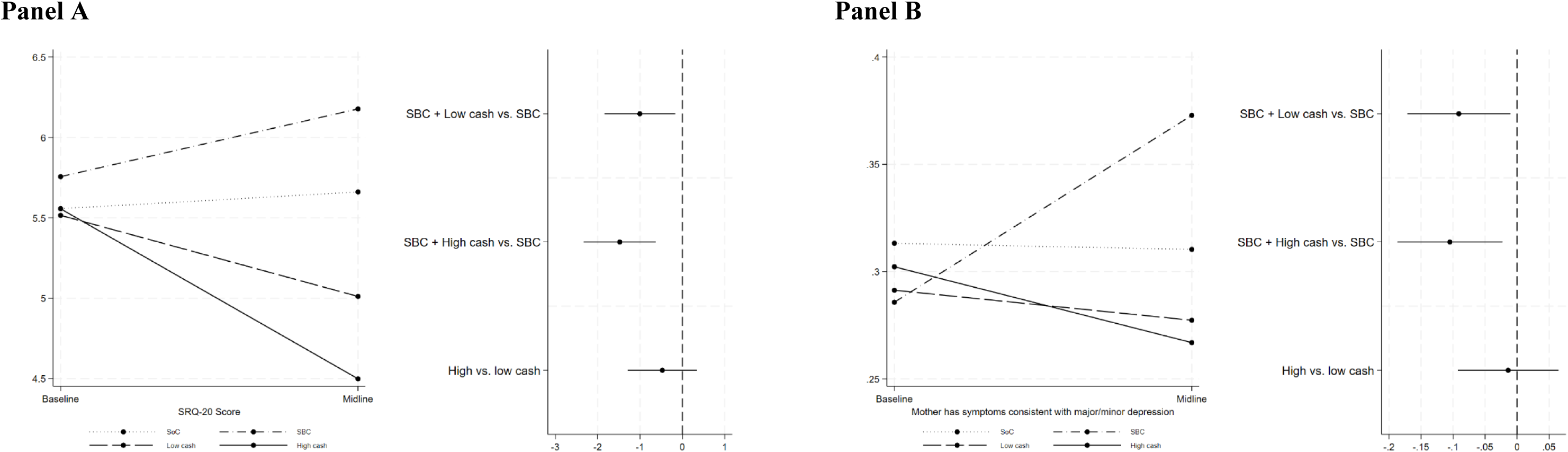
Added benefit of the low and high cash transfers in addition to the nutrition-sensitive social behaviour change interventions on depression scores (Panel A) and the proportion of women with depressive symptoms (Panel B). Abbreviations used: SBC, social behaviour change.

We found limited evidence of effect modification of the effect of the MAZIKO interventions on SRQ-20 scores or the proportion of women with depressive symptoms (**Supplemental Tables 7and 8**). Specifically, child sex modified the effect of the SBC+high cash intervention on the proportion of women with depressive symptoms (p-vale for interaction <0.10) with larger effects for mothers with boys and not girls (Supplemental Table 8).

## Discussion

This study assessed the impact of cash transfers integrated with nutrition-sensitive SBC interventions on perinatal mental health during the lean season in Malawi. Eighteen months post-baseline, the SBC+high cash interventions reduced depression scores as compared to SoC, with reductions corresponding to an effect size of -0.27 SMD. The SBC+high cash interventions benefited all women, not just those with more severe depressive symptoms. The SBC+low cash and SBC alone interventions had no impact on maternal mental health relative to SoC. Our study also demonstrated that those women who were provided with cash transfers with SBC in comparison to just SBC had lower depression scores regardless of the size of the cash transfer. Reductions corresponded to an effect size of -0.24 SMD for SBC+low cash vs SBC and -0.35 SMD for SBC+high cash vs SBC. When comparing the SBC+high cash to SBC+low cash groups, there was no evidence of added benefit of the larger cash transfer relative to the smaller cash transfer. In addition, we found that mothers of boys appeared to benefit more from the SBC+high cash intervention than mothers of girls.

Our study demonstrated that the most comprehensive package (combining SoC, SBC, and large cash transfers) was the most effective relative to the SoC in improving maternal perinatal depression. We hypothesised that the interventions worked by addressing the highest number of vulnerabilities and stressors, and supported the largest variety of positive coping and thriving strategies during the lean season. Post-endline analyses will unpack the specific pathways through which the interventions worked to improve maternal mental health.^51^ The effect size of -0.27 SMD we observed was nearly 4 times higher than the effect size of -0.07 SMD of parenting interventions alone^40^ and 2.7 times higher than the effect size of -0.10 SMD of cash transfers alone.^23^ Taken together, these estimates support the hypothesis that cash transfers, nutrition-sensitive agriculture, and parenting interventions can have additive or synergistic effects on maternal mental health during periods of high food insecurity.

Further, the size of the cash transfer appeared to matter in generating additive or synergistic effects. Although depressive symptoms declined in the SBC+low cash arm, we found no impact on depressive symptoms of the SBC+low cash interventions relative to SoC. These findings are consistent with a study in Tanzania which found no impact of combining small cash transfers (TZS5,000-10,000 or USD2.20-4.30 per visit) with a parenting intervention on perinatal mental health^29^ They are also consistent with the social protection literature indicating larger effects on mental health with larger cash transfers.^20–23^ Likewise, we found that SBC alone had no impact on mental health relative to SoC. In fact, depression scores increased in the SBC arm. This may be because of the low self-reported participation in SBC activities. Although anecdotal evidence indicated misreporting of participation for fear of losing programme benefits, it is more likely that women limited their participation in more labour or effort intensive activities in the last trimester of pregnancy and first few months post-partum. Moreover, most of the benefits from the agriculture and livelihoods activities (the only SBC interventions fully rolled out by midline) are likely not immediate, constant every month, or predictable like the cash transfers. Thus, there were likely insufficient activities to promote and protect women’s mental health or benefits that had yet to be generated. Yet another explanation is that without cash women had insufficient resources to act on the advice received through SBC,^58^ which may have increased their stress.

That SBC+low cash or SBC alone were insufficient to help women and their households cope with existing or new vulnerabilities is not surprising given the many shocks in the country in the months prior to the midline survey. In 2023, Malawi experienced nearly 30% inflation.^59^ In March 2023, tropical cyclone Freddy displaced 659,000 people and caused loss of homes and livelihoods for many others.^60^ El Niño induced a late onset of the 2023/2024 rainy season, inadequate rains, and reduced the number of rainy days negatively affecting crop production.^61^ Humanitarian emergencies increase risks to mental health not only by disrupting livelihoods and financial/economic stability, but also by disrupting health services, increasing food insecurity, poverty, overcrowding, and breakdown of social networks.^1^ Considering all these events, many respondents in the MAZIKO midline qualitative evaluation, conducted in April-June 2024, reported feeling worried and stressed because of their inability to provide sufficient and diverse food for their families.^58^ Another MAZIKO article showed that while SBC+high cash protected household consumption and reduced food insecurity at midline, there were no effects in the SBC+low cash or SBC arms.^62^ These results support our hypothesis that the SBC and SBC+low cash were insufficient to address economic vulnerabilities and reduce food insecurity during the lean season following major economic and climate shocks. Notably, only the agriculture and livelihoods components of the SBC were fully rolled out by midline. Had the Caring for the Caregiver and Male Champions interventions been rolled out earlier, larger and more diverse impacts on maternal perinatal depression may have been observed. The late roll out for both components was necessary to align activities with the government’s wider piloting and training plans for these two interventions.

Relative to SBC, both cash amounts improved women’s mental health. Although the effect size was larger for the SBC+high cash than for the SBC+low cash interventions (-0.23 and -0.35 SMD, respectively), the difference was not statistically significant. Notably, these effect sizes are similar to the -0.27 SMD effect size for the effect of SBC+high cash vs SoC. Coupled with the increase in depression scores in the SBC arm, these findings indicate that the positive impacts on maternal perinatal depression are likely driven by the cash transfers. However, given the integrated nature of the interventions, we cannot fully disentangle which intervention components contributed to the improvement in mental health, and we cannot rule out additive or synergistic effects.

Our study is among a handful to evaluate the effect of cash transfers on perinatal depression and the first to evaluate the effect of combining cash transfers with nutrition-sensitive SBC (comprising agriculture and livelihoods, male engagement, and psychosocial interventions) on perinatal depression. The interventions were assessed through a rigorous cluster-randomised controlled trial, relying on a theory-driven approach. Since the study was implemented in a high number of clusters in two districts, results are generalisable to the rest of the country. Nevertheless, some limitations of this work should be noted. First, we only surveyed half the sample at midline, prioritising women who self-reported as pregnant at baseline. As a result, our findings may not generalise to the rest of the sample, consisting of women with children 1.5-3.5 years of age at the time of the midline survey. Whereas 80% of women in the midline sample were still breastfeeding, most of the women in the remainder of the baseline sample would no longer be breastfeeding and may likely have been more worried and stressed about providing food for their children. Thus, it is unclear whether the SBC+high cash interventions would have been sufficient to improve mental health for these women. Second, the trial did not include a pure control group or cash only arms. Since the government SoC was available to all pregnant women and mothers of young children, including a pure control group was infeasible. We did not include cash only arms because we preferred to maximise the number of new comparisons, considering the extensive social protection literature on the impacts of cash transfers alone on variety of household and individual outcomes.

## Conclusion

Overall, larger cash transfers coupled with nutrition-sensitive SBC interventions comprising nutrition-sensitive agriculture, male engagement, and psychosocial components were effective in improving maternal perinatal depression during the lean season, benefiting all women not just those with more severe depressive symptoms. Cash transfers provided an added benefit on top of SBC. The interventions worked by addressing multiple vulnerabilities and risks and providing a range of coping and thriving strategies. Targeted cash transfers and SBC interventions during the key perinatal period can help promote and protect mental health during peak periods of food insecurity. In low-income countries like Malawi, there are <1 mental health workers per 100,000 population,^1^ and substantial financial and human resources investments are needed before the health system can implement other cost-effective interventions for mental health.^63^ Nevertheless, the SBC package is comprehensive and both values of the cash transfer are large relative to other cash transfer programs in Malawi or programmes targeting perinatal mental health. Planned cost-effectiveness analyses at endline^51^ will further build the evidence base for cost-effective community-based interventions for perinatal mental health and guide next steps in optimising the scale-up of the MAZIKO interventions, and in designing new programmes to improve women’s wellbeing.

## Contributors

Aulo Gelli led the MAZIKO trial design with contributions from Melissa Gladstone, Mangani Katundu, Kenneth Maleta, and Natalie Roschnik. Lilia Bliznashka, Marilyn Ahun, Aulo Gelli, Melissa Gladstone, Mangani Katundu, and Kenneth Maleta wrote the trial protocol. Brenda Phiri and Esnatt Gondwe-Matekesa supervised intervention implementation. Lilia Bliznashka and Aulo Gelli conceptualised the analyses presented here. Peter Mvula, Alister

Munthali, and Monice Kachinjika implemented the surveys and supervised data collection. Lilia Bliznashka, Odiche Nwabuikwu, and Daniel Maggio oversaw training of data collection. Lilia Bliznashka and Odiche Nwabuikwu performed data cleaning, curation, and analysis. Lilia Bliznashka drafted the manuscript. All authors interpreted the results and reviewed and approved the final version of the manuscript. Lilia Bliznashka had final responsibility for submitting the manuscript for publication.

## Patient consent for publication

Not applicable.

## Ethical considerations

The trial was approved by the institutional review boards of the University of Malawi (protocol number: 02/22/128) and the International Food Policy Research Institute (protocol number: PHND-22-0214).

## Provenance and peer review

Not commissioned; externally peer reviewed.

## Data sharing

Data described in the manuscript, code book, and analytic code will be made available by the corresponding author upon request.

## Acknowledgements

We would like to thank enumerators and respondents for their time and willingness to participate in the study.

## The MAZIKO trial team

### Analysis and writing committee

Aulo Gelli (Principal Investigator), Jan Duchoslav, Melissa Gladstone, Daniel Gilligan, Mangani Katundu, Kenneth Maleta, Agnes Quisumbing, Lilia Bliznashka, Marilyn Ahun.

### Programme team

At Save the Children: Burcu Munyas, Brenda Phiri, Anthony Kulemba, Olita Mulewa, Takondwa Minjale, Sarah Chasowa, Chisoni Lundu, Judith Mnyawa, Deusdedit Dambuleni. Supporting staff: Jean Nkhonjera, Phindile Lupafya, Felix Mtonda, Chimwewe Kamala, Steve Kamtimaleka, Frank Mwafulirwa, Rachel Dixon, George Chidalengwa, John Chipeta, Thandizolathu Kadzamira, Ashebir Debebe, Raquel Tebaldi, Shelagh Possmayer, Stephen Mutiso.

At Give Directly: Shaunak Ganguly, Yvonne Namala Murindiwa, Esnatt Gondwe-Matekesa, Dingaan Kafundu, Chisomo Banda, Bhahart Msuku, Ireen Kanjala.

### Doctoral student collaborators

Karoline Becker, Anissa Collishaw, Daniel Maggio, Natalie Roschnik, Aisha Twalibu.

### Collaborators

Carol Levin, Cheryl Doss, Dan Maggio^1^.

### Field Management team

Peter Mvula, Monice Kachinjika, Theresa Nnensa, Alister Munthali, Victoria Ndolo.

### Data Management Odiche Nwabuikwu. Trial steering group

IFPRI: Marie Ruel, Harold Alderman. Save the Children: Joanne Grace. Give Directly: Miriam Laker

^1^Following graduation from doctoral program in 2023.

## References

1. WHO. World mental health report: transforming mental health for all. Geneva: World Health Organization, 2022.

2 Bluett-Duncan M, Kishore MT, Patil DM, Satyanarayana VA, Sharp H. A systematic review of the association between perinatal depression and cognitive development in infancy in low and middle-income countries. PLoS One 2021; 16: e0253790.

3 Morales MF, Girard L-C, Raouna A, MacBeth A. The association of different presentations of maternal depression with children’s socio-emotional development: A systematic review. PLOS Glob Public Heal 2023; 3: e0001649.

4 Carosella E, Chhabria S, Kim H, et al. Perinatal depression and adverse child growth outcomes in low-income and middle-income countries (LMICs): A systematic review and meta-analysis. PLOS Glob Public Heal 2024; 4: e0003586.

5 Gelaye B, Rondon MB, Araya R, Williams MA. Epidemiology of maternal depression, risk factors, and child outcomes in low-income and middle-income countries. The Lancet Psychiatry 2016; 3: 973–82.

6 Accortt EE, Cheadle ACD, Dunkel Schetter C. Prenatal Depression and Adverse Birth Outcomes: An Updated Systematic Review. Matern Child Health J 2015; 19: 1306–37.

7 Rahman A, Fisher J, Bower P, et al. Interventions for common perinatal mental disorders in women in low- and middle-income countries: a systematic review and meta-analysis. Bull World Health Organ 2013; 91: 593–601I.

8 Slomian J, Honvo G, Emonts P, Reginster J-Y, Bruyère O. Consequences of maternal postpartum depression: A systematic review of maternal and infant outcomes. Women’s Heal 2019; 15. DOI:10.1177/1745506519844044.

9 McLearn KT, Minkovitz CS, Strobino DM, Marks E, Hou W. The Timing of Maternal Depressive Symptoms and Mothers’ Parenting Practices With Young Children: Implications for Pediatric Practice. Pediatrics 2006; 118: e174–82.

10. IHME. GBD Compare. 2021. https://vizhub.healthdata.org/gbd-compare/# (accessed Sept 7, 2024).

11 Chorwe-Sungani G, Wella K, Mapulanga P, Nyirongo D, Pindani M. Systematic review on the prevalence of perinatal depression in Malawi. South African J Psychiatry 2022; 28. DOI:10.4102/sajpsychiatry.v28i0.1859.

12 Chorwe-Sungani G, Chipps J. A cross-sectional study of depression among women attending antenatal clinics in Blantyre district, Malawi. South African J Psychiatry 2018; 24. DOI:10.4102/sajpsychiatry.v24i0.1181.

13 Stewart RC, Bunn J, Vokhiwa M, et al. Common mental disorder and associated factors amongst women with young infants in rural Malawi. Soc Psychiatry Psychiatr Epidemiol 2010; 45: 551–9.

14 Stewart RC, Umar E, Tomenson B, Creed F. A cross-sectional study of antenatal depression and associated factors in Malawi. Arch Womens Ment Health 2014; 17: 145–54.

15 Stewart RC, Umar E, Gleadow-Ware S, Creed F, Bristow K. Perinatal distress and depression in Malawi: an exploratory qualitative study of stressors, supports and symptoms. Arch Womens Ment Health 2015; 18: 177–85.

16 LeMasters K, Dussault J, Barrington C, et al. ‘Pain in my heart’: Understanding perinatal depression among women living with HIV in Malawi. PLoS One 2020; 15: e0227935.

17 Patel V, Saxena S, Lund C, et al. The Lancet Commission on global mental health and sustainable development. Lancet 2018; 392: 1553–98.

18 Lund C, Brooke-Sumner C, Baingana F, et al. Social determinants of mental disorders and the Sustainable Development Goals: a systematic review of reviews. The Lancet Psychiatry 2018; 5: 357–69.

19 Ridley M, Rao G, Schilbach F, Patel V. Poverty, depression, and anxiety: Causal evidence and mechanisms. Science (80-) 2020; 370. DOI:10.1126/science.aay0214.

20 McGuire J, Kaiser C, Bach-Mortensen AM. A systematic review and meta-analysis of the impact of cash transfers on subjective well-being and mental health in low- and middle-income countries. Nat Hum Behav 2022; 6: 359–70.

21 Gomboc S, Zagoranski M, Kos A, et al. Systematic Review on the Impact of Various Types of Universal Basic Income on Mental Health in Low- and Middle-Income Countries. Behav Sci (Basel) 2024; 14: 726.

22 Oswald TK, Nguyen MT, Mirza L, et al. Interventions targeting social determinants of mental disorders and the Sustainable Development Goals: a systematic review of reviews. Psychol Med 2024; 54: 1475–99.

23 Wollburg C, Steinert JI, Reeves A, Nye E. Do cash transfers alleviate common mental disorders in low- and middle-income countries? A systematic review and meta-analysis. PLoS One 2023; 18: e0281283.

24 Kilburn K, Handa S, Angeles G, Tsoka M, Mvula P. Paying for Happiness: Experimental Results from a Large Cash Transfer Program in Malawi. J Policy Anal Manag 2018; 37: 331–56.

25 Angeles G, de Hoop J, Handa S, Kilburn K, Milazzo A, Peterman A. Government of Malawi’s unconditional cash transfer improves youth mental health. Soc Sci Med 2019; 225: 108–19.

26 Molotsky A, Handa S. The Psychology of Poverty: Evidence from the Field. J Afr Econ 2021; 30: 207–24.

27 Ohrnberger J, Fichera E, Sutton M, Anselmi L. The worse the better? Quantile treatment effects of a conditional cash transfer programme on mental health. Health Policy Plan 2020; 35: 1137–49.

28 Powell-Jackson T, Pereira SK, Dutt V, Tougher S, Haldar K, Kumar P. Cash transfers, maternal depression and emotional well-being: Quasi-experimental evidence from India’s Janani Suraksha Yojana programme. Soc Sci Med 2016; 162: 210–8.

29 Bliznashka L, Yousafzai AK, Asheri G, Masanja H, Sudfeld CR. Effects of a community health worker delivered intervention on maternal depressive symptoms in rural Tanzania. Health Policy Plan 2020; published online Dec 13. DOI:10.1093/heapol/czaa170.

30. FAO. Food Security. 2006. https://www.fao.org/fileadmin/templates/faoitaly/documents/pdf/pdf_Food_Security_Cocept_Note.pdf.

31 Weaver LJ, Hadley C. Moving Beyond Hunger and Nutrition: A Systematic Review of the Evidence Linking Food Insecurity and Mental Health in Developing Countries. Ecol Food Nutr 2009; 48: 263–84.

32 Sparling TM, Nesbitt RC, Henschke N, Gabrysch S. Nutrients and perinatal depression: a systematic review. J Nutr Sci 2017; 6: e61.

33 Pourmotabbed A, Moradi S, Babaei A, et al. Food insecurity and mental health: a systematic review and meta-analysis. Public Health Nutr 2020; 23: 1778–90.

34 Ruel MT, Quisumbing AR, Balagamwala M. Nutrition-sensitive agriculture: What have we learned so far? Glob Food Sec 2018; : 1–26.

35 Rosenberg AM, Maluccio JA, Harris J, et al. Nutrition-sensitive agricultural interventions, agricultural diversity, food access and child dietary diversity: Evidence from rural Zambia. Food Policy 2018; 80: 10–23.

36 Kangmennaang J, Kerr RB, Lupafya E, Dakishoni L, Katundu M, Luginaah I. Impact of a participatory agroecological development project on household wealth and food security in Malawi. Food Secur 2017; 9: 561–76.

37 Cetrone HM, Santoso M V, Bezner Kerr R, et al. Food security mediates the decrease in women’s depressive symptoms in a participatory nutrition-sensitive agroecology intervention in rural Tanzania. Public Health Nutr 2021; 24: 4682–92.

38 Santoso M V, Bezner Kerr RN, Kassim N, et al. A Nutrition-Sensitive Agroecology Intervention in Rural Tanzania Increases Children’s Dietary Diversity and Household Food Security But Does Not Change Child Anthropometry: Results from a Cluster-Randomized Trial. J Nutr 2021; published online May 11. DOI:10.1093/jn/nxab052.

39. World Health Organization. Mental health: strengthening our response. 2018. https://www.who.int/news-room/fact-sheets/detail/mental-health-strengthening-our-response (accessed Sept 7, 2020).

40 Jeong J, Franchett EE, Ramos de Oliveira C V., Rehmani K, Yousafzai AK. Parenting interventions to promote early child development in the first three years of life: A global systematic review and meta-analysis. PLOS Med 2021; 18: e1003602.

41 Jeong J, Pitchik HO, Yousafzai AK. Stimulation Interventions and Parenting in Low- and Middle-Income Countries: A Meta-analysis. Pediatrics 2018; 141: e20173510.

42 Al Sager A, Goodman SH, Jeong J, Bain PA, Ahun MN. Effects of multi-component parenting and parental mental health interventions on early childhood development and parent outcomes: a systematic review and meta-analysis. Lancet Child Adolesc Heal 2024; 8: 656–69.

43 Singla DR, Kumbakumba E, Aboud FE. Effects of a parenting intervention to address maternal psychological wellbeing and child development and growth in rural Uganda: A community-based, cluster-randomised trial. Lancet Glob Heal 2015; 3: e458–69.

44 Atukunda P, Muhoozi GKM, Westerberg AC, Iversen PO. Nutrition, Hygiene and Stimulation Education for Impoverished Mothers in Rural Uganda: Effect on Maternal Depression Symptoms and Their Associations to Child Development Outcomes. Nutrients 2019; 11: 1561.

45 Rockers PC, Zanolini A, Banda B, et al. Two-year impact of community-based health screening and parenting groups on child development in Zambia: Follow-up to a cluster-randomized controlled trial. PLOS Med 2018; 15: e1002555.

46 Jeong J, Sullivan EF, McCann JK. Effectiveness of father-inclusive interventions on maternal, paternal, couples, and early child outcomes in low- and middle-income countries: A systematic review. Soc Sci Med 2023; 328: 115971.

47 Clarke K, King M, Prost A. Psychosocial Interventions for Perinatal Common Mental Disorders Delivered by Providers Who Are Not Mental Health Specialists in Low- and Middle-Income Countries: A Systematic Review and Meta-Analysis. PLoS Med 2013; 10: e1001541.

48 Singla DR, Kohrt BA, Murray LK, Anand A, Chorpita BF, Patel V. Psychological Treatments for the World: Lessons from Low- and Middle-Income Countries. Annu Rev Clin Psychol 2017; 13: 149–81.

49 Gelli A, Aberman N-L, Margolies A, Santacroce M, Baulch B, Chirwa E. Lean-Season Food Transfers Affect Children’s Diets and Household Food Security: Evidence from a Quasi-Experiment in Malawi. J Nutr 2017; 147: 869–78.

50 Devereux S, Vaitla B, Swan SH. Seasons of Hunger. Pluto Press, 2015 DOI:10.2307/j.ctt183q3rs.

51 Gelli A, Duchoslav J, Gladstone M, et al. Impact evaluation of a maternal and child cash transfer intervention, integrated with nutrition, early childhood development, and agriculture messaging (MAZIKO-IE): a study protocol for a cluster-randomised controlled trial. Trials 2024; 25: 46.

52. Save the Children, Give Directly, IFPRI. MAZIKO - Malawi integrated maternal and child grant project. 2023. https://resourcecentre.savethechildren.net/pdf/MAZIKO-Malawi-Integrated-Maternal-and-Child-Grant-Project.pdf/.

53 Bliznashka L, Nwabuikwu O, Ahun M, et al. Understanding modifiable caregiver factors contributing to child development among young children in rural Malawi. Matern Child Nutr 2024; published online July 3. DOI:10.1111/mcn.13698.

54 Becker K, Bliznashka L, Doss C, et al. Associations between women’s empowerment and maternal depressive symptoms: A cross-sectional analysis from Balaka and Ntcheu districts in Malawi. 2023 DOI:10.2499/p15738coll2.137078.

55 Leight J, Pedehombga A, Ganaba R, Gelli A. Women’s empowerment, maternal depression, and stress: Evidence from rural Burkina Faso. SSM - Ment Heal 2022; 2: 100160.

56 World Health Organization. A user’s guide to the self-reporting questionnaire (SRQ). Geneva, 1994.

57 Stewart RC, Kauye F, Umar E, et al. Validation of a Chichewa version of the Self-Reporting Questionnaire (SRQ) as a brief screening measure for maternal depressive disorder in Malawi, Africa. J Affect Disord 2009; 112: 126–34.

58. Bath SRD. Cash Plus for Nutrition and Child Development in Malawi: MAZIKO midterm Qualitative Impact Protocol (QuIP) evaluation. 2024 https://resourcecentre.savethechildren.net/pdf/Maziko-QuIP-report-Final-_14_11_24b.pdf/.

59. IMF. Inflation rate, average consumer prices. IMF Datamapper. 2024. https://www.imf.org/external/datamapper/PCPIPCH@WEO/MWI?year=2024 (accessed Nov 15, 2024).

60. Internal Displacement Monitoring Center. Malawi - Cyclone Freddy puts disaster risk management to the test. 2024 Glob. Rep. Intern. Displac. 2024. https://www.internal-displacement.org/spotlights/Malawi-Cyclone-Freddy-puts-disaster-risk-management-to-the-test/ (accessed Nov 15, 2024).

61. United Nations Malawi. National El Niño Induced Prolonged Dry Spells and Floods Response Appeal. 2024. https://malawi.un.org/en/273587-national-el-niño-induced-prolonged-dry-spells-and-floods-response-appeal (accessed Nov 15, 2024).

62 Collishaw A, Maggio D, Gelli A, Maziko Trial Team. Nutrition-Sensitive Cash Transfers and Lean Season Food Insecurity. https://ace.illinois.edu/sites/default/files/2024-11/JMP Collishaw 11-2024.pdf.

63. WHO. WHO Menu of cost-effective interventions for mental health. Geneva: World Health Organization, 2021.

