## Supplemental for "Effect of combining lower- and higher-value monthly cash transfers with nutrition-sensitive agriculture, male engagement, and psychosocial intervention on maternal depressive symptoms in rural Malawi: a secondary analysis of a cluster-randomised controlled trial"

**Supplemental Table 1** Proportion of women reporting each symptom on the Self-Reporting Questionnaire (SRQ-20) at baseline and midline by intervention arm

| Symptom | Survey round | Standard of care | Social behaviour change (SBC) | SBC + low cash | SBC + high cash |
|---|---|---|---|---|---|
| Often has headaches | Baseline | 55.7 | 59.6 | 57.4 | 54.0 |
| | Midline | 48.3 | 56.4 | 51.8 | 48.2 |
| Poor appetite | Baseline | 26.7 | 28.2 | 29.7 | 26.7 |
| | Midline | 18.7 | 19.2 | 22.4 | 18.0 |
| Poor sleep | Baseline | 36.8 | 36.6 | 36.4 | 36.7 |
| | Midline | 29.0 | 33.8 | 25.5 | 28.0 |
| Shaking hands | Baseline | 11.8 | 15.7 | 15.1 | 13.8 |
| | Midline | 14.9 | 10.5 | 12.9 | 10.9 |
| Feeling nervous, tense or worried | Baseline | 40.2 | 48.1 | 41.2 | 38.9 |
| | Midline | 53.7 | 55.4 | 43.4 | 42.1 |
| Easily frightened | Baseline | 20.1 | 20.6 | 21.8 | 19.3 |
| | Midline | 25.6 | 25.1 | 21.6 | 20.3 |
| Poor digestion | Baseline | 7.8 | 7.0 | 8.7 | 5.1 |
| | Midline | 4.3 | 7.3 | 4.5 | 3.2 |
| Has trouble thinking clearly | Baseline | 21.8 | 25.1 | 24.6 | 23.8 |
| | Midline | 31.0 | 33.4 | 27.7 | 24.8 |
| Feeling unhappy | Baseline | 32.5 | 36.9 | 32.8 | 33.1 |
| | Midline | 37.6 | 43.2 | 30.5 | 28.6 |
| Cries more than usual | Baseline | 9.5 | 7.7 | 8.4 | 6.4 |
| | Midline | 9.5 | 12.2 | 7.6 | 5.8 |
| Hard to enjoy daily activities | Baseline | 20.4 | 19.9 | 20.4 | 18.0 |
| | Midline | 25.9 | 34.1 | 22.7 | 18.0 |
| Hard to make decisions | Baseline | 13.8 | 18.1 | 15.4 | 19.0 |
| | Midline | 24.4 | 30.0 | 23.5 | 18.6 |
| Daily work is suffering | Baseline | 23.0 | 24.0 | 20.7 | 23.8 |
| | Midline | 27.3 | 31.7 | 25.5 | 23.2 |
| Cannot play a useful part in life | Baseline | 28.7 | 28.9 | 27.2 | 27.0 |
| | Midline | 38.2 | 36.6 | 32.8 | 25.4 |
| Lost interest in things | Baseline | 22.1 | 24.7 | 18.8 | 23.2 |
| | Midline | 28.4 | 25.8 | 22.7 | 19.6 |
| Feeling worthless | Baseline | 24.4 | 19.2 | 19.6 | 20.9 |
| | Midline | 30.7 | 34.5 | 24.9 | 23.2 |
| Thoughts of ending one's life | Baseline | 9.5 | 7.3 | 6.7 | 6.4 |
| | Midline | 13.2 | 9.8 | 6.2 | 5.5 |
| Feeling tired all the time | Baseline | 47.4 | 44.9 | 48.5 | 49.8 |

| | | | | | |
|---|---|---|---|---|---|
| | Midline | 32.2 | 38.3 | 28.3 | 29.3 |
| Uncomfortable feelings in one's stomach | Baseline | 48.6 | 48.4 | 43.7 | 50.5 |
| | Midline | 36.8 | 42.9 | 34.7 | 31.2 |
| Easily tired | Baseline | 54.9 | 54.7 | 54.3 | 59.2 |
| | Midline | 36.2 | 37.6 | 31.9 | 26.0 |

**Supplemental Table 2** Unadjusted effects of the MAZIKO interventions relative to the standard of care

| | Baseline | | | | Midline | | | | Unadjusted estimates | | |
|---|---|---|---|---|---|---|---|---|---|---|---|
| | SOC Mean ± SD or % | SBC Mean ± SD or % | SBC + Low cash Mean ± SD or % | SBC + High cash Mean ± SD or % | SOC Mean ± SD or % | SBC Mean ± SD or % | SBC + Low cash Mean ± SD or % | SBC + High cash Mean ± SD or % | SBC vs SOC Mean difference (95% CI) | SBC + Low cash vs SOC Mean difference (95% CI) | SBC + High cash vs SOC Mean difference (95% CI) |
| N | 348 | 287 | 357 | 311 | 348 | 287 | 357 | 311 | 1303 | 1303 | 1303 |
| Self-Reporting Questionnaire (SRQ-20) score | 5.6±4.2 | 5.8±4.2 | 5.5±4.3 | 5.6±4.1 | 5.7±4.8 | 6.2±4.6 | 5.0±4.6 | 4.5±4.3 | 0.34 (-0.49, 1.18) | -0.66 (-1.47, 0.15) | -1.13 (-1.96, -0.31) |
| Mother has symptoms consistent with depression (SRQ-20≥8) | 31.3 | 28.6 | 29.1 | 30.2 | 31.0 | 37.3 | 27.7 | 26.7 | 0.06 (-0.02, 0.14) | -0.03 (-0.11, 0.05) | -0.04 (-0.12, 0.04) |

Abbreviations used: SBC, social behaviour change; SOC, standard of care.

**Supplemental Table 3** Adjusted effect of the MAZIKO interventions relative to the standard of care

| | **Baseline** | | | | **Midline** | | | | **Adjusted estimates** | | |
|---|---|---|---|---|---|---|---|---|---|---|---|
| | **SOC Mean ± SD or %** | **SBC Mean ± SD or %** | **SBC + Low cash Mean ± SD or %** | **SBC + High cash Mean ± SD or %** | **SOC Mean ± SD or %** | **SBC Mean ± SD or %** | **SBC + Low cash Mean ± SD or %** | **SBC + High cash Mean ± SD or %** | **SBC vs SOC Mean difference (95% CI)** | **SBC + Low cash vs SOC Mean difference (95% CI)** | **SBC + High cash vs SOC Mean difference (95% CI)** |
| N | 348 | 287 | 357 | 311 | 348 | 287 | 357 | 311 | 1303 | 1303 | 1303 |
| Self-Reporting Questionnaire (SRQ-20) score | 5.6±4.2 | 5.8±4.2 | 5.5±4.3 | 5.6±4.1 | 5.7±4.8 | 6.2±4.6 | 5.0±4.6 | 4.5±4.3 | 0.41 (-0.38, 1.19) | -0.59 (-1.34, 0.16) | -1.08 (-1.84, -0.31) |
| Mother has symptoms consistent with depression (SRQ-20≥8) | 31.3 | 28.6 | 29.1 | 30.2 | 31.0 | 37.3 | 27.7 | 26.7 | 0.07 (-0.01, 0.15) | -0.02 (-0.10, 0.05) | -0.04 (-0.11, 0.04) |

Estimates adjusted for district and baseline covariates: woman's age (<18 vs ≥18 y), education (primary/lower vs secondar/higher), marital status (married vs unmarried), household head status (self vs other), household head gender (female vs male), and household wealth (lowest vs higher quintiles of total household expenditures). Abbreviations used: SBC, social behaviour change; SOC, standard of care.

**Supplemental Table 4** Unadjusted effect of the MAZIKO interventions relative to the standard of care excluding 50 women who did not receive cash transfers

| | **Baseline** | | | | **Midline** | | | | **Unadjusted estimates** | | |
|---|---|---|---|---|---|---|---|---|---|---|---|
| | **SOC Mean ± SD or %** | **SBC Mean ± SD or %** | **SBC + Low cash Mean ± SD or %** | **SBC + High cash Mean ± SD or %** | **SOC Mean ± SD or %** | **SBC Mean ± SD or %** | **SBC + Low cash Mean ± SD or %** | **SBC + High cash Mean ± SD or %** | **SBC vs SOC Mean difference (95% CI)** | **SBC + Low cash vs SOC Mean difference (95% CI)** | **SBC + High cash vs SOC Mean difference (95% CI)** |
| N | 348 | 287 | 334 | 284 | 348 | 287 | 334 | 284 | 1253 | 1253 | 1253 |
| Self-Reporting Questionnaire (SRQ-20) score | 5.6±4.2 | 5.8±4.2 | 5.4±4.3 | 5.5±4.1 | 5.7±4.8 | 6.2±4.6 | 4.8±4.5 | 4.4±4.3 | 0.33 (-0.53, 1.2) | -0.79 (-1.63, 0.05) | -1.17 (-2.03, -0.31) |
| Mother has symptoms consistent with depression (SRQ-20≥8) | 31.3 | 28.6 | 29.0 | 29.9 | 31.0 | 37.3 | 26.1 | 25.7 | 0.06 (-0.02, 0.14) | -0.04 (-0.12, 0.04) | -0.05 (-0.13, 0.03) |

Abbreviations used: SBC, social behaviour change; SOC, standard of care.

**Supplemental Table 5** Added benefit of the low and high cash transfers in addition to the nutrition-sensitive social behaviour change interventions (unadjusted effects)

| | Baseline | | | | Midline | | | | Unadjusted estimates | | |
|---|---|---|---|---|---|---|---|---|---|---|---|
| | SOC Mean ± SD or % | SBC Mean ± SD or % | SBC + Low cash Mean ± SD or % | SBC + High cash Mean ± SD or % | SOC Mean ± SD or % | SBC Mean ± SD or % | SBC + Low cash Mean ± SD or % | SBC + High cash Mean ± SD or % | SBC + Low cash vs SBC Mean difference (95% CI) | SBC + How cash vs SBC Mean difference (95% CI) | SBC + High cash vs SBC + low cash Mean difference (95% CI) |
| N | 348 | 287 | 357 | 311 | 348 | 287 | 357 | 311 | 1303 | 1303 | 1303 |
| Self-Reporting Questionnaire (SRQ-20) score | 5.6±4.2 | 5.8±4.2 | 5.5±4.3 | 5.6±4.1 | 5.7±4.8 | 6.2±4.6 | 5.0±4.6 | 4.5±4.3 | -1.00 (-1.84, -0.17) | -1.48 (-2.33, -0.63) | -0.47 (-1.29, 0.35) |
| Mother has symptoms consistent with depression (SRQ-20≥8) | 31.3 | 28.6 | 29.1 | 30.2 | 31.0 | 37.3 | 27.7 | 26.7 | -0.09 (-0.17, -0.01) | -0.11 (-0.19, -0.02) | -0.01 (-0.09, 0.06) |

Abbreviations used: SBC, social behaviour change.

**Supplemental Table 6** Added benefit of the low and high cash transfers in addition to the nutrition-sensitive social behaviour change interventions (unadjusted effects)

| | Baseline | | | | Midline | | | | Adjusted estimates | | |
|---|---|---|---|---|---|---|---|---|---|---|---|
| | **SOC Mean ± SD or %** | **SBC Mean ± SD or %** | **SBC + Low cash Mean ± SD or %** | **SBC + High cash Mean ± SD or %** | **SOC Mean ± SD or %** | **SBC Mean ± SD or %** | **SBC + Low cash Mean ± SD or %** | **SBC + High cash Mean ± SD or %** | **SBC + Low cash vs SBC Mean difference (95% CI)** | **SBC + How cash vs SBC Mean difference (95% CI)** | **SBC + High cash vs SBC + low cash Mean difference (95% CI)** |
| N | 348 | 287 | 357 | 311 | 348 | 287 | 357 | 311 | 1303 | 1303 | 1303 |
| Self-Reporting Questionnaire (SRQ-20) score | 5.6±4.2 | 5.8±4.2 | 5.5±4.3 | 5.6±4.1 | 5.7±4.8 | 6.2±4.6 | 5.0±4.6 | 4.5±4.3 | -1.00 (-1.78, -0.22) | -1.49 (-2.28, -0.69) | -0.48 (-1.25, 0.28) |
| Mother has symptoms consistent with depression (SRQ-20≥8) | 31.3 | 28.6 | 29.1 | 30.2 | 31.0 | 37.3 | 27.7 | 26.7 | -0.09 (-0.17, -0.01) | -0.11 (-0.18, -0.03) | -0.01 (-0.09, 0.06) |

Estimates adjusted for district, and baseline covariates: woman's age (<18 vs ≥18 y), education (primary/lower vs secondar/higher), marital status (married vs unmarried), household head status (self vs other), household head gender (female vs male), and household wealth (lowest vs higher quintiles of total household expenditures). Abbreviations used: SBC, social behaviour change.

**Supplemental Table 7** Modification of the effect of the MAZIKO interventions on depression scores.

| | SBC vs. SOC | SBC + Low cash vs. SOC | SBC + High cash vs. SOC |
|---|---|---|---|
| | **Mean Difference (95% CI)** | **Mean Difference (95% CI)** | **Mean Difference (95% CI)** |
| Child is a girl | 0.14 (-0.97, 1.26) | -0.85 (-1.9, 0.2) | -1.12 (-2.18, -0.06) |
| Child is a boy | 0.35 (-0.76, 1.46) | -0.58 (-1.64, 0.48) | -1.6 (-2.71, -0.49) |
| p-value | 0.77 | 0.68 | 0.48 |
| Household is in higher expenditure quintiles | 0.17 (-0.72, 1.07) | -0.73 (-1.6, 0.13) | -1.13 (-2.01, -0.24) |
| Household is in lowest expenditure quintile | 1.26 (-0.34, 2.86) | -0.32 (-1.79, 1.15) | -1.15 (-2.62, 0.33) |
| p-value | 0.20 | 0.61 | 0.98 |
| Not pregnant at baseline | 0.52 (-1.54, 2.58) | -1.28 (-3.17, 0.61) | -0.92 (-2.98, 1.14) |
| Pregnant at baseline | 0.3 (-0.57, 1.18) | -0.64 (-1.49, 0.2) | -1.22 (-2.08, -0.36) |
| p-value | 0.84 | 0.52 | 0.78 |

Abbreviations used: SBC, social behaviour change; SOC, standard of care.

**Supplemental Table 8** Modification of the effect of the MAZIKO interventions on the proportion of women with depressive symptoms.

| | **SBC vs. SOC** | **SBC + Low cash vs. SOC** | **SBC + High cash vs. SOC** |
|---|---|---|---|
| | **Mean Difference (95% CI)** | **Mean Difference (95% CI)** | **Mean Difference (95% CI)** |
| Child is a girl | 0.05 (-0.06, 0.16) | -0.02 (-0.13, 0.08) | -0.01 (-0.12, 0.09) |
| Child is a boy | 0.05 (-0.07, 0.16) | -0.06 (-0.17, 0.04) | -0.14 (-0.26, -0.03) |
| p-value | 0.94 | 0.56 | 0.08 |
| Household is in higher expenditure quintiles | 0.06 (-0.03, 0.14) | -0.03 (-0.12, 0.05) | -0.05 (-0.14, 0.04) |
| Household is in lowest expenditure quintile | 0.11 (-0.05, 0.28) | 0.01 (-0.14, 0.16) | -0.01 (-0.16, 0.14) |
| p-value | 0.52 | 0.62 | 0.61 |
| Not pregnant at baseline | 0.04 (-0.17, 0.26) | -0.12 (-0.32, 0.07) | -0.05 (-0.26, 0.16) |
| Pregnant at baseline | 0.06 (-0.02, 0.15) | -0.02 (-0.1, 0.06) | -0.05 (-0.13, 0.03) |
| p-value | 0.85 | 0.31 | 0.99 |

Abbreviations used: SBC, social behaviour change; SOC, standard of care.

**Supplemental Figure 1** Distribution of SRQ-20 scores by intervention arm and survey

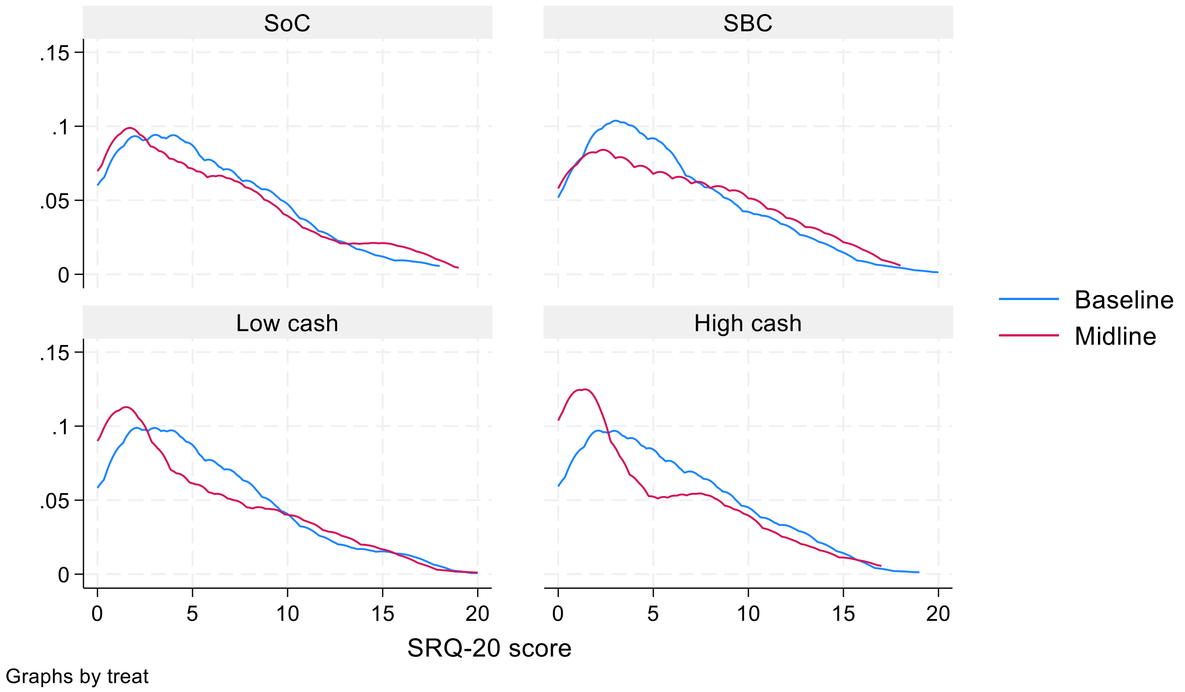
